# An International, Cross-Sectional Survey of Psychiatry Researchers and Clinicians: Perceptions of Complementary, Alternative, and Integrative Medicine

**DOI:** 10.1101/2024.01.24.24301718

**Authors:** Jeremy Y. Ng, Jassimar Kochhar, Holger Cramer

## Abstract

**Background:** Psychiatry is a branch of medicine that focuses on mental, behavioural and emotional well-being. Complementary, alternative, and integrative medicine has been an increasingly popular choice for patients with psychiatric disorders, therefore our study aimed to explore the perceptions of psychiatry researchers and clinicians on the use of CAIM.

**Methods:** We conducted an online, anonymous, cross-sectional survey for researchers and clinicians who have published their work in psychiatry medical journals that are indexed in MEDLINE. 42,667 researchers and clinicians were sent the link to the survey after extraction of their email addresses from their respective publications. Respondents were asked numerous multiple-choice questions regarding their perceptions on various CAIM therapies, followed by an open-ended question where they could include any additional thoughts.

**Results:** The survey was completed by 987 respondents, with a majority identifying as a researcher (n=447, 46.51%), or as both a researcher and a clinician (n=368, 38.29%) within the field of psychiatry. Most respondents (n=629, 78.04%) perceived mind-body therapies such as meditation, biofeedback, hypnosis, and yoga to be the most promising CAIM therapy for the prevention, treatment, and management of psychiatric diseases. Many participants said that they agree (n=285, 38.14%) that most CAIM therapies in general are safe, however, many disagree that CAIM therapies are effective (n=245, 32.93%). Respondents indicated that there is value to conducting research on CAIM therapies (n=356, 47.91%), and that there should be more funding allocated to researching these therapies (n=265, 35.71%). Respondents are also in agreement that clinicians should receive training on CAIM therapies through formal education (n=295, 39.76%) or supplementary education (n=380, 51.28%).

**Conclusions:** The findings from this study showed that there is great interest and potential in researching CAIM within the field of psychiatry. This information can be used as a basis for further research and to develop tailored educational resources for researchers and clinicians in psychiatry.

## Background

Psychiatry is a medical specialty that encompasses mental, emotional, and behavioural issues [1]. The term “psychiatric disorder” covers a wide range of difficulties that may disturb an individual’s mood, behaviour, thoughts, or feelings [1]. These disorders are also often associated with impaired functioning [1] and can have a significant impact on the individual’s daily life [2]. Specific diagnostic criteria for respective psychiatric conditions are found in the Diagnostic and Statistical Manual of Mental Disorders-5 (DSM-5) [3], and the International Classification of Disease (ICD-11) [4]. The most prevalent psychiatric disorders are anxiety and depressive disorders, with the number rising, 26% and 28%, respectively, since the emergence of the COVID-19 pandemic [5]. Symptoms of psychiatric disorders vary depending on the condition and severity [3]; however, many individuals will suffer with mood fluctuations, reduced concentration, inability to cope with stress, changes in eating habits, and an increase in suicidal thinking [5]. Individuals with severe mental health conditions also have shorter life expectancies [6], demonstrating that psychiatric disorders continue to be a leading cause of disability and mortality. Globally, 1 in every 8 people are currently living with a psychiatric or mental disorder [5]. Therefore, it is imperative to determine effective treatment options in dealing with this mental health crisis. Patients that suffer with psychiatric disorders have been increasingly considering complementary, alternative, and integrative medicine (CAIM), especially over the past 20 years to combat their symptoms, or to counter side effects that they face with other treatments [7]. In the past, it has been reported that 9.8% of individuals who have a mental health disorder such as anxiety, mood and alcohol disorders have utilized CAIM [8]. “Complementary medicine” refers to practices that are not considered mainstream which are used in conjunction with conventional medicine. Conversely, “alternative medicine” is classified by a non-mainstream approach being used in lieu of conventional medicine. Finally, “integrative health” refers to the coordinated use of both conventional and complementary approaches [9–10]. For the purpose of this study, we will collectively refer to this group of diverse therapies as CAIM.

There are a wide range of CAIM therapies with varying degrees of benefits. Certain CAIMs may have beneficial effects in treating psychiatric conditions such as depression and anxiety, especially when used in conjunction with conventional treatment [11–12]. This includes S-adenosylmethionine (SAM-e), St. John’s wort, omega-3 fatty acids, folate, and exercise [13]. SAM-e has been found to be implicated in depression, as patients typically have low serum and cerebral spinal fluid levels of this naturally occurring molecule. Supplementation of SAM-e raises levels of dopamine and other beneficial neurotransmitters [13]. St. John’s wort is one of the most investigated CAIM therapies, and it is hypothesized to inhibit synaptosomal uptake of monoamine neurotransmitters [14]. However, it is important to consider that it is a contraindication to several other pharmaceutical agents such as human immunodeficiency virus (HIV) protease inhibitors, immunosuppressants, hormonal contraceptives, serotonergic agents and other antidepressants [15]. While the role that omega-3 fatty acids play in improving depressive disorders is not fully understood, it has been found that patients with depression have lower serum levels of omega-3, suggesting supplementation may be beneficial in relieving symptoms [16]. Folate is another essential molecule involved in the synthesis of the aforementioned neurotransmitters [13]. Finally, the role of exercise in improving psychiatric conditions is widely researched, with studies showing its ability to alter the function of neurotransmitters, and lower cortisol levels [13].

The existing literature [17–18] demonstrates that many patients have utilized or would like to further explore the use of CAIM to relieve their symptoms, however, it has also been found that many healthcare practitioners do not receive adequate training and education about CAIM [19], making it difficult for them to appropriately recommend these therapies to patients [20]. In addition, clinical practice guideline recommendations for a range of CAIMs pertaining to the treatment and management of disorders such as depression and anxiety are uncertain, as these recommendations contain great variability and are of sub-optimal quality [21–22]. Thus, it would be valuable to understand the current perceptions towards CAIM among psychiatry researchers and clinicians. A better understanding on the views of psychiatry researchers and clinicians towards CAIM therapies may allow for the development of tailored educational resources, and therefore improve knowledge on the topic. Consequently, this can lead to better integration of evidence-based CAIM therapies in clinical settings, and a move away from non-evidence-based practices – [19], and therefore better communication between healthcare practitioner and the patient, and eventually improve patient outcomes [23]. The purpose of this study is therefore to determine the perceptions of psychiatry researchers and clinicians about complementary, alternative, and integrative medicine.

## Methods

### Transparency Statement

Before commencing this project, clearance and approval from the University Hospital Tübingen Research Ethics Board (REB) was obtained (REB Number: 389/2023BO2). Additionally, the study was preregistered on the Open Science Framework, and the study protocol is available at <u>10.17605/OSF.IO/75ACV</u>. The final copy of the manuscript will be posted on preprinted prior to submission to a journal.

### Study Design

This study was conducted as an online, anonymous, cross-sectional survey for researchers and clinicians who have published their work in psychiatry medical journals that were indexed in MEDLINE.

### Sampling Framework

The selection occurred from a sample of authors who have published articles in psychiatry journals found at https://www.ncbi.nlm.nih.gov/nlmcatalog?term=Psychiatry%5Bst%5D over the time period April 1, 2017 to May 1, 2023. The search strategy included published records of any type. The NLM IDs of the journals were first extracted from the results of an OVID MEDLINE search strategy of articles published in psychiatry journals (see: https://osf.io/wz75p). To retrieve the authors’ name, affiliation institutions, and email addresses, the list of PMIDs that were found by the search were exported as .csv files. There were 62 of these .csv files, which contained 2000 records each. These were then placed in a master sheet of PMIDs, and 200 PMIDs at a time were uploaded onto R studio using the “easyPubMed” and “readr” packages.

### Participant Recruitment

The sampling framework was utilized to create a list of contact information consisting of individuals who were likely to be mostly psychiatry researchers and clinicians, as they have published their articles in the psychiatry journals indexed in MEDLINE. These individuals were contacted to complete the survey and participate in the study. The platform that was used to create the survey and send the emails was Survey Monkey [24]. A pre-written email approved by the REB containing details regarding the goals of the study as well as a link to the survey was sent to the prospective participants in this email. Once participants clicked the survey link, they were taken to the first page of the survey on SurveyMonkey. At the bottom of this page, participants were asked if they consented to the aforementioned terms and conditions associated with participating in the survey, and must have answered “Yes” to this question to proceed to view the questions of the survey. Before sending the recruitment emails, duplicate addresses were removed from the dataset. The data also underwent a process of cleaning to fix names that were extracted incorrectly, such as names with accents which contain incorrect characters, and to remove names that did not have email addresses. To encourage responses, participants were sent reminder emails after the first, second, and third weeks from the initial email. The participants had four weeks to complete the survey after their third and final reminder, and if they encountered any questions they did not wish to respond to, they were able to skip them. The survey was open from August 21, 2023 until September 18, 2023.

### Survey Design

Participants were first asked a screening question, which was then followed by demographic questions. The remaining questions asked participants about their perceptions of CAIM. The format of most of the questions were multiple choice (quantitative data), with one open-ended question (qualitative data) at the end, and it is anticipated that the survey took approximately 15 minutes to complete. Two independent CAIM researchers pilot tested the survey before it was distributed. A copy of the survey can be found on the following link: https://osf.io/5t4bx.

### Data Management and Analysis

The responses to the multiple-choice questions were used to generate basic descriptive statistics such as counts and percentages. Additionally, in order to analyze the qualitative data, a thematic content analysis was conducted [25]. Specifically, the responses from the open-ended questions were interpreted and assigned a code, which was a short representation of their response.

## Results

### Demographics

This search strategy yielded 121,460 articles in total, from which there were 122,307 PMIDs. The contact information of 705,179 individuals was then extracted. After the removal of duplicates, special characters, and misspelt names, the final list contained 42,676 unique email addresses. There were 42, 667 emails sent, of which 20, 719 were unopened, and 5772 were bounced. Overall, this survey had 987 responses (2.7% response rate of unopened and opened, and 6.1% of just the opened). Raw survey data (with any personal identifiers redacted) are available at: https://osf.io/9jwpn. It is also important to note there were a select few participants who did not respond to all the questions. If no questions were answered after the initial screening question, the responses were deemed incomplete. The average time spent to complete the survey was 9 minutes. The majority of respondents in the study indicated that they identify as a researcher (n=447, 46.51%), or as both a researcher and a clinician (n=368, 38.29%) within the field of psychiatry. The most common World Health Organization World Regions of the respondents included Americas (n=353, 40.90%), Europe (n=325, 37.66%), and South-East Asia (n=78, 9.04%). Most respondents indicated that they were faculty members/principal investigators (n=484, 56.02%), and working as a senior researcher or clinician with >10 years of starting their career post formal education (n=497, 57.72%). Additionally, a large proportion of participants responded that their primary research area is in clinical research (n=530, 70.20%), and epidemiological research (n=225, 29.80%). Further information regarding the survey participant characteristics can be found in **Table 1**.

**Table 1:** Characteristics of Survey Participants.

|  |  |
| --- | --- |
| **Sex (n=861)** | n= (%) |
| Male | 418 (48.55%) |
| Female | 424 (49.25%) |
| Intersex | 0 (0.00%) |
| Prefer not to say | 15 (1.74%) |
| Prefer to self describe | 4 (0.46%) |
| **Age (n=861)** |  |
| Under 18 | 0 (0.00%) |
| 18-24 | 1 (0.12%) |
| 25-34 | 138 (16.03%) |
| 35-44 | 252 (29.27%) |
| 45-54 | 209 (24.27%) |
| 55-64 | 135 (15.68%) |
| 65 or older | 114 (13.24%) |
| Prefer not to say | 12 (1.39%) |
| **Visible Minority (n=862)** |  |
| Yes | 112 (12.99%) |
| No | 720 (83.53%) |
| Prefer not to say | 30 (3.48%) |
| **World Health Organization World Region (n=863)** |  |
| Africa | 13 (1.51%) |
| Americas | 353 (40.90%) |
| Eastern Mediterranean | 25 (2.90%) |
| Europe | 325 (37.66%) |
| South-East Asia | 78 (9.04%) |
| Western Pacific | 56 (6.49%) |
| Prefer not to say | 13 (1.51%) |
| **Current Position (n=864)** |  |
| Clinician Student | 16 (1.85%) |
| Clinician | 270 (31.25%) |
| Graduate student | 37 (4.28%) |
| Postdoctoral fellow | 93 (10.76%) |
| Faculty member/principal investigator | 484 (56.02%) |
| Research support staff | 35 (4.05%) |
| Scientist in academia | 270 (31.25%) |
| Scientist in industry | 9 (1.04%) |
| Scientist in third sector | 19 (2.20%) |
| Government scientist | 24 (2.78%) |
| Other | 27 (3.13%) |
| **Career Stage (n=861)** |  |
| Graduate or clinician student | 30 (3.48%) |
| Early career researcher or clinician (<5 years post education) | 135 (15.68%) |
| Mid-career researcher or clinician (5-10 years post education) | 199 (23.11%) |
| Senior researcher or clinician (>10 years post education) | 497 (57.72%) |
| **Primary research area (n=755)** |  |
| Clinical research | 530 (70.20%) |
| Preclinical research – in vivo | 92 (12.19%) |
| Preclinical research – in vitro | 34 (4.50%) |
| Health systems research | 94 (12.45%) |
| Health services research | 181 (23.97%) |
| Methods research | 104 (13.77%) |
| Epidemiological research | 225 (29.80%) |
| Other | 52 (6.89%) |

### Complementary, Alternative, and Integrative Medicine

There was a high number of respondents (n=446, 59.47%) who have never conducted any CAIM research, however majority of them (n=629, 78.04%) perceived mind-body therapies such as meditation, biofeedback, hypnosis, and yoga to be the most promising CAIM therapy for the prevention, treatment, and management of psychiatric diseases/conditions (**Figure 1**). In addition, mind-body therapy was indicated to be the one which most of their patients had sought counselling or disclosed using (n=340, 85.64%), with biologically based practices such as vitamins and dietary supplements being the next (n=302, 76.07%). When asked what percentage of patients disclosed using CAIM or seek counselling on therapies over the past year, most respondents answered a rate of only 0-10% (n=120, 30.85%). Mind-body therapies (n=276, 69.52%) and biologically based practices (n=145, 36.52%) were also the most common therapies that respondents practiced or recommended to patients. More than half of respondents had not received any formal training in any areas of CAIM (n=214, 54.45%), however a large number of respondents received supplemental training in mind-body therapies (n=193, 48.74%). Most researchers (n=758, 93.81%) used academic literature or conference presentations/workshops to seek more information on CAIM. Outside of the research or clinical setting, many respondents (n=489, 60.44%) indicated that they have been asked about CAIM occasionally. When asked if they believe that most CAIM therapies in general are safe, many participants said that they agree (n=285, 38.14%), However, when asked if CAIM therapies are effective, many disagreed (n=245, 32.93%). Many respondents believed that there is value to conducting research on CAIM therapies (n=356, 47.91%), and that there should be more research funding allocated to study these therapies (n=265, 35.71%). Respondents also agreed that clinicians should receive training on CAIM therapies via formal education (n=295, 39.76%) or via supplementary education (n=380, 51.28%) (**Figure 2**). A very similar numbers of respondents answered agree (n=105, 28.93%) and disagree (n=106, 29.20%) when asked if they would be comfortable counselling patients about CAIM therapies, however approximately one-third (n=125, 34.44%) disagreed that they would be comfortable recommending most CAIM therapies to their patients.

**Figure 1.**
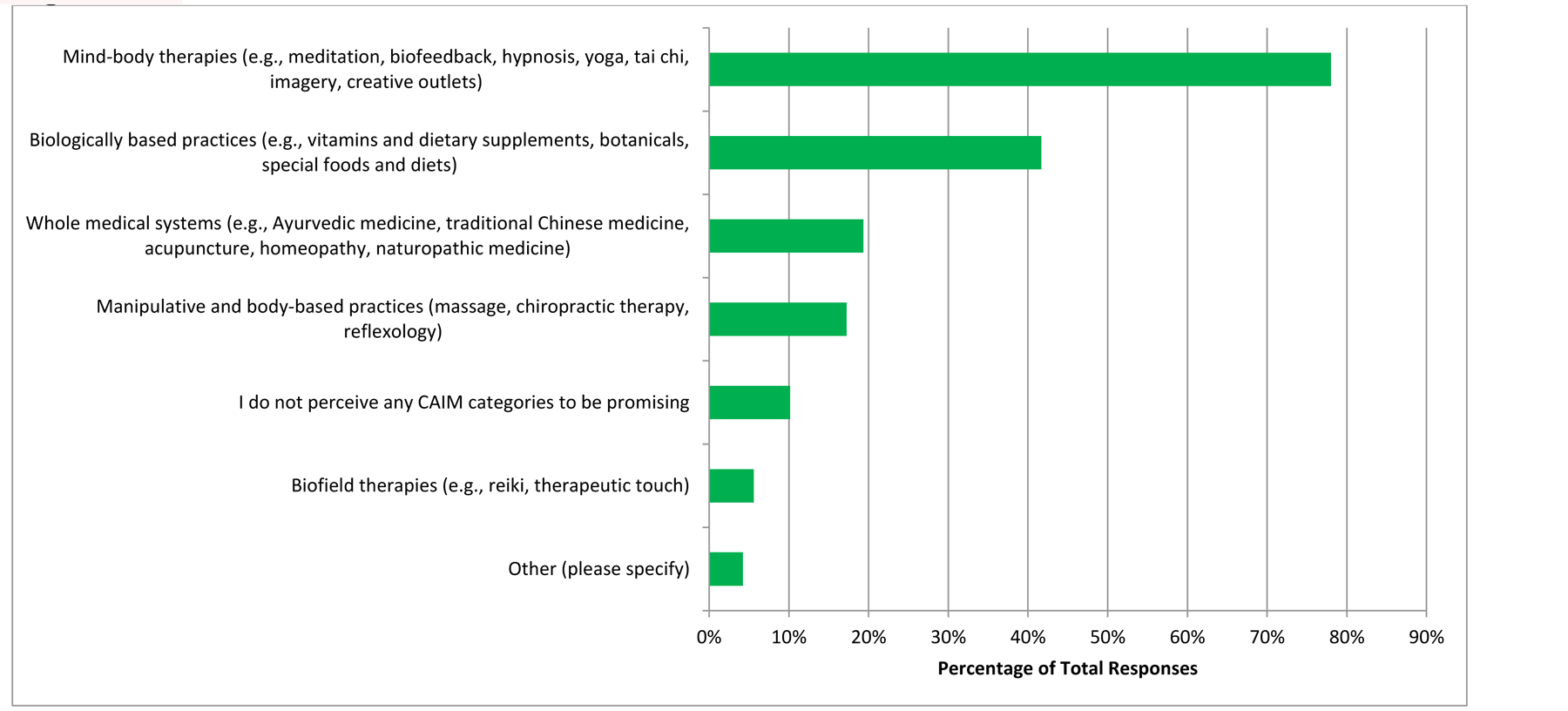
CAIM Category Perceived to be the Most Promising in Psychiatry.

**Figure 2.**
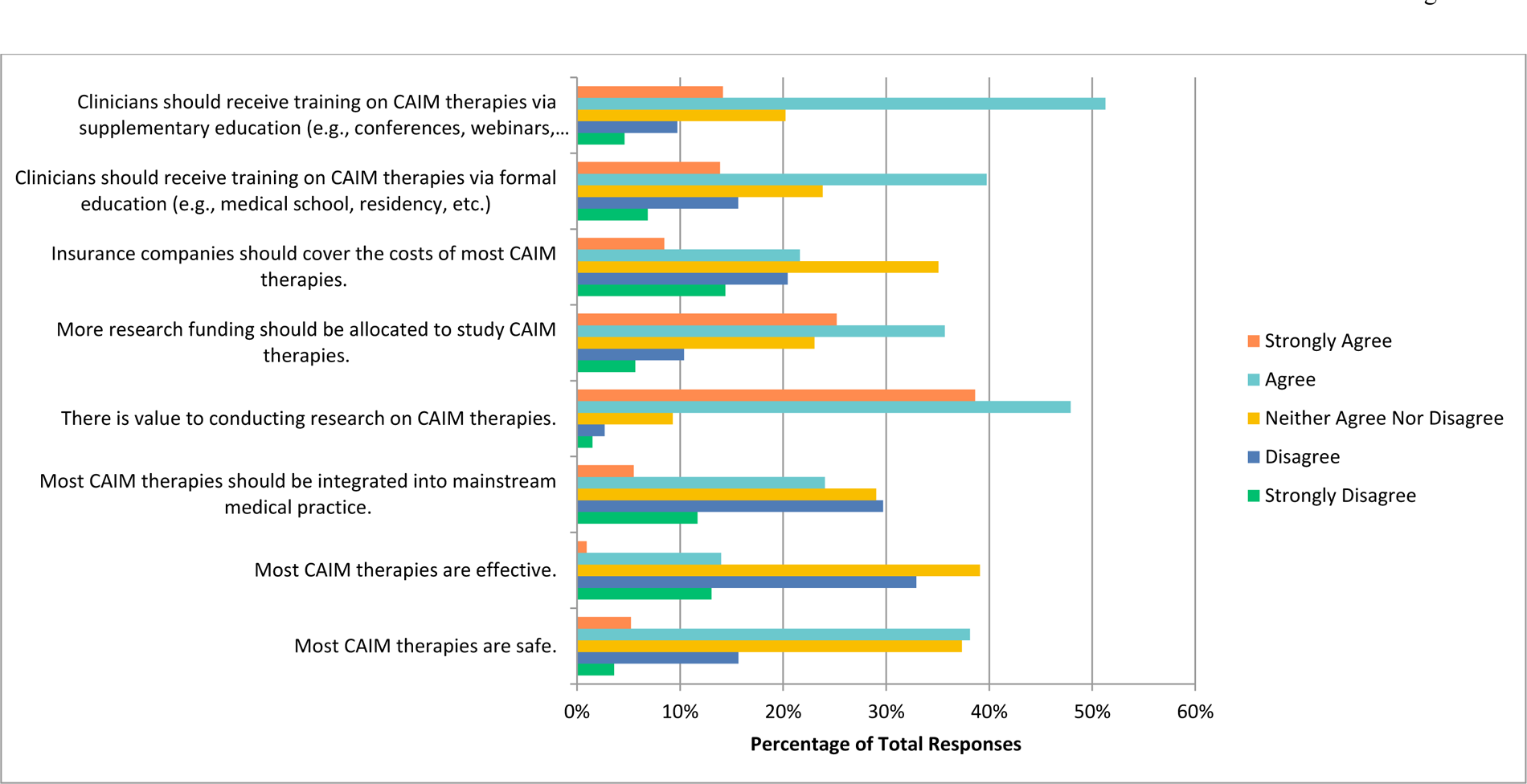
Agreement with the Following Statements Regarding CAIM in General.

### Mind Body Therapy

When asked about mind-body therapies in particular, majority of respondents (n=57.76%) agreed it is safe, that it should be integrated into mainstream medical practices (n=282, 38.21%), that there is value to conducting research on this topic (n=357, 48.24%), that more research funding should be allocated towards this study (n=306, 41.46%), and finally that clinicians should receive training on mind-body therapies via formal education (n=319, 43.22%) or supplemental education (n=403, 54.76%) (**Figure 3**). Majority of them also agreed that they would be comfortable counselling patients about most mind-body therapies (n=171, 47.50%), and recommending most of them to their patients (n=134, 37.22%).

**Figure 3.**
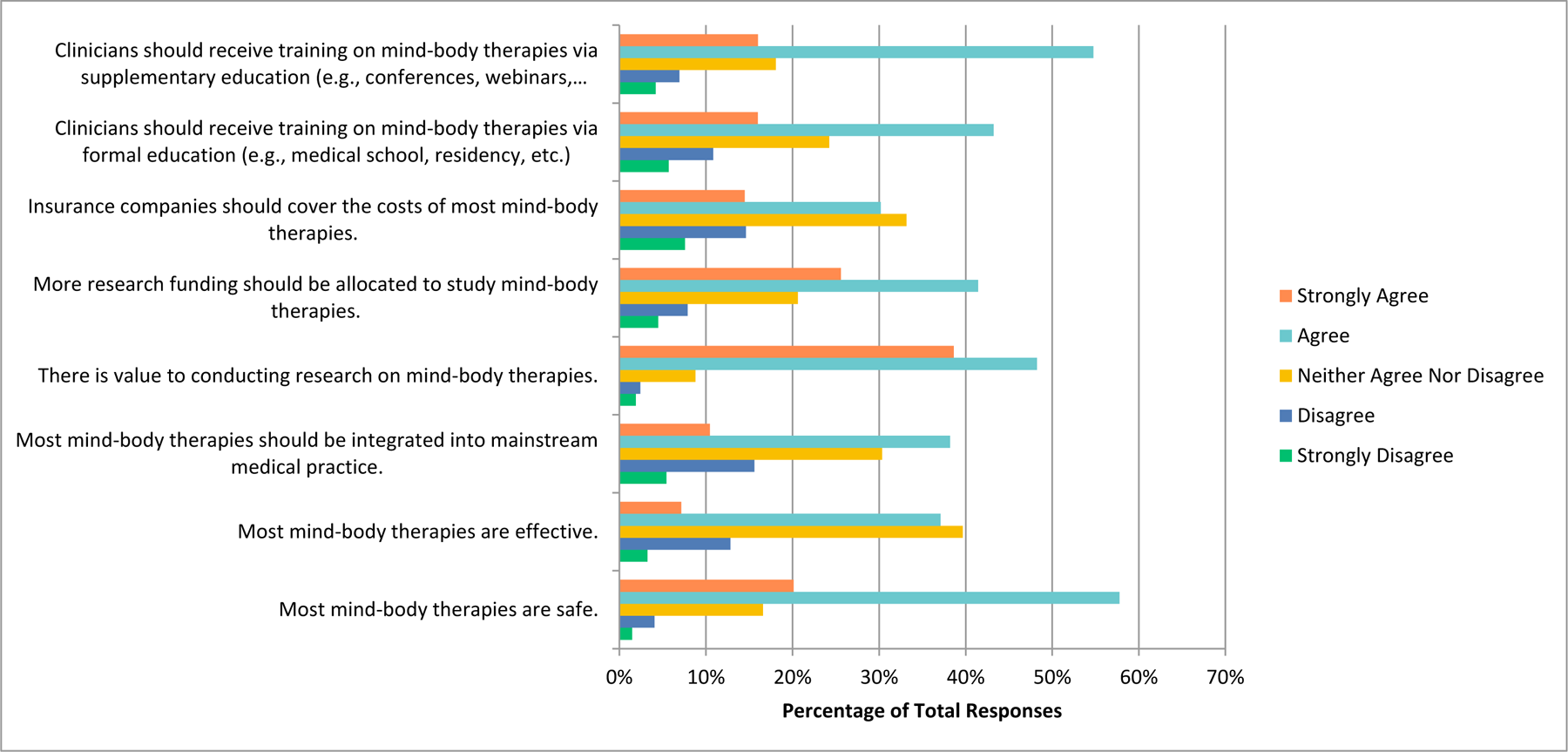
Agreement with the Following Statements Regarding Mind-Body Therapies.

### Biologically Based Practices

In relation to biologically based practices, respondents agreed that there is value to conducting research on this topic (n=395, 54.26%), that more research funding should be allocated towards it (n=277, 38%), that clinicians should receive training via formal education (n=297, 40.74%) and supplemental education (n=334, 45.94%). However, many respondents seemed unsure and answered that they neither agreed nor disagreed when asked if these practices are safe (n=313, 42.94%), effective (n=311, 42.72%), should be integrated into mainstream medicine (n=270, 37.14%), and that insurance should cover their costs (n=279, 38.27%) (**Figure 4**). Most respondents agreed that they would be comfortable counselling their patients about these therapies (n=108, 30.08%), however most disagreed that they would be comfortable in recommending these therapies to patients (n=102, 28.49%).

**Figure 4.**
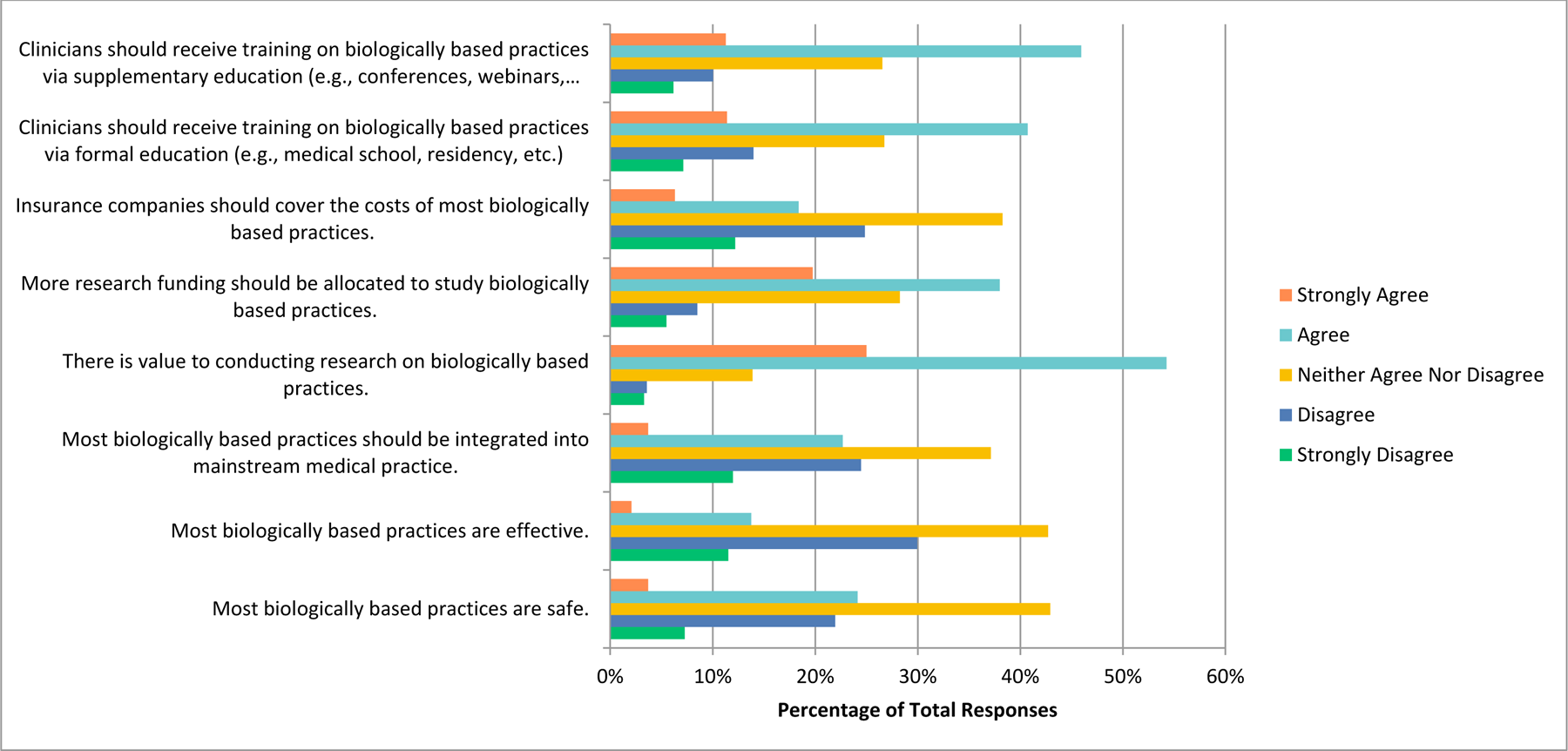
Agreement with the Following Statements Regarding Biologically Based Practices.

### Manipulative and Body-Based Practices

The manipulative and body-based practices question mostly received responses indicating that participants neither agreed nor disagreed when asked if they are safe (n=311, 42.60%), effective (n=333, 45.62%), should be integrated into medicine (n=302, 41.37%), funds should be allocated to study this (n=254, 34.84%), insurance companies should cover their costs (n=300, 41.10%) and that clinicians should receive training via formal education (n=279, 38.17%) or via supplementary education (n=257, 35.35%) (**Figure 5**). Respondents also answered that they neither agreed nor disagreed when asked if they would be comfortable counselling patients about this therapy (n=105, 29.25%) and if they would be comfortable recommending this therapy to patients (n=106, 29.53%). However, majority agreed that there is value to conducting research in this area (n=344, 47.12%).

**Figure 5.**
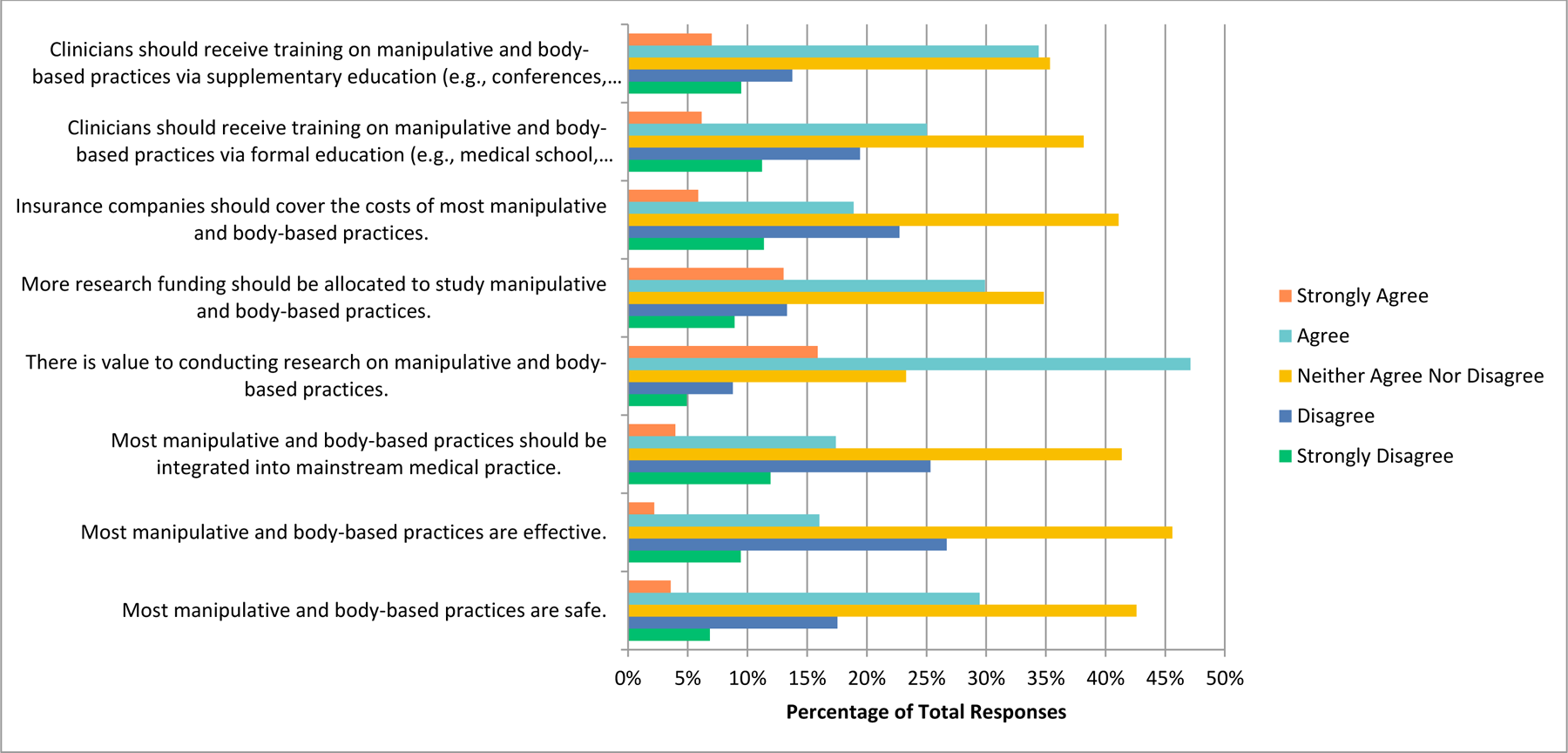
Agreement with the Following Statements Regarding Manipulative and Body-Based Practices.

### Biofield Therapies

The respondents answered similarly when asked about biofield therapies, as they neither agreed nor disagreed when asked if they believed these are safe (n=369, 50.83%), effective (n= 293,40.41%), should be integrated into mainstream medical practice (n=263,36. 23%), that there is value to conducting research in this field (n=219, 30.12%), that more research funding should be allocated to study this area (n=247, 33.98%), that insurance companies should cover the costs (n=260, 35.81%), and that clinicians should receive training via formal education (n=253, 34.85%) or supplementary education (n=260, 35.91%) (**Figure 6**). Respondents also strongly disagreed when asked if they would be comfortable counselling patients about these therapies (n=117, 32.68%), or recommending therapies to patients (n=159, 44.54%).

**Figure 6.**
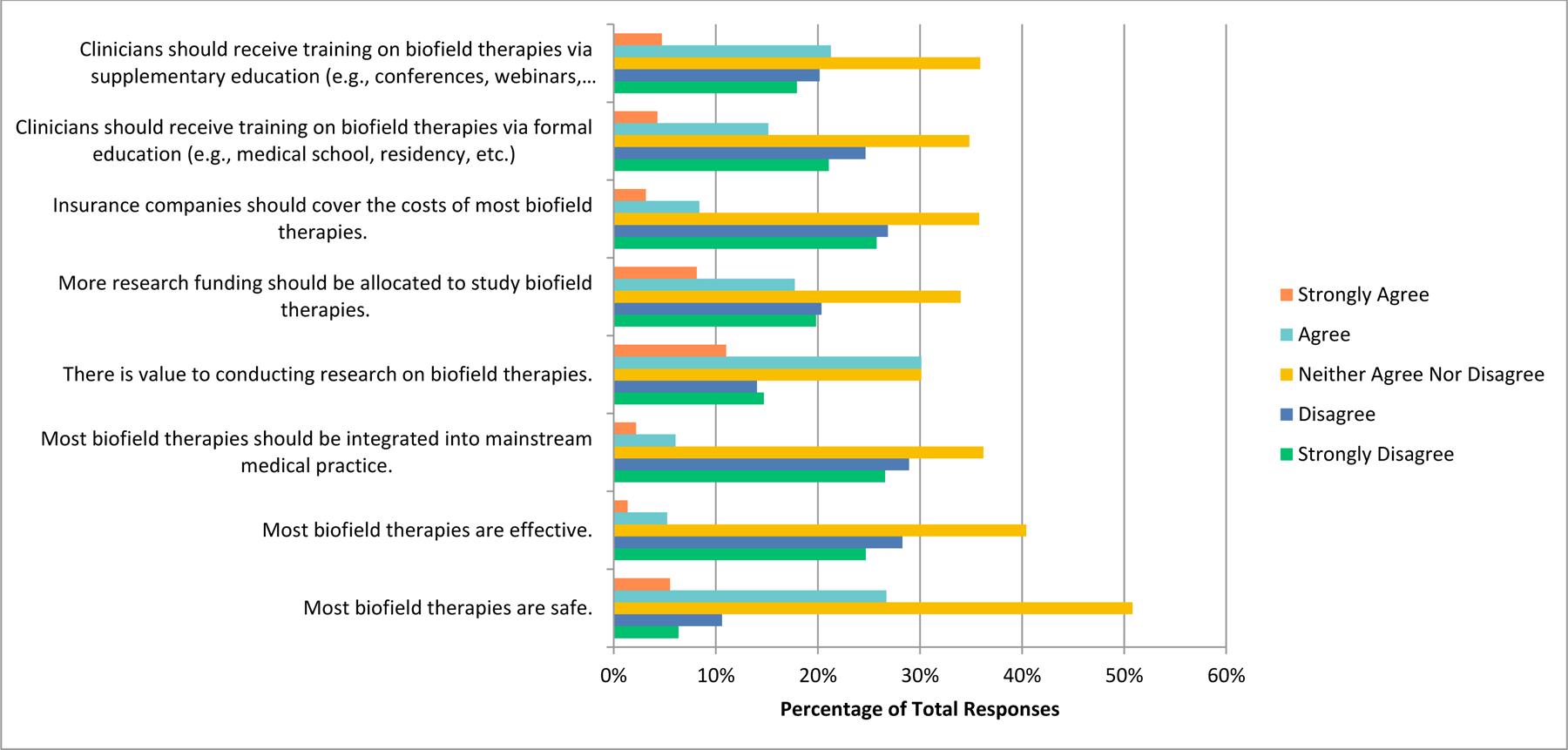
Agreement with the Following Statements Regarding Biofield Therapies.

### Whole Medical Systems

In regards to whole medical systems, many respondents neither agreed nor disagreed when asked if these therapies are safe (n=314, 43.07%), effective (n=314, 43.19%), should be integrated into mainstream medical practices (n=256, 35.16%), that insurance should cover the costs of this service (n= 273, 37.55%), and that clinicians should receive training via formal education (n=235, 32.28%). However, they also answered that they agreed when asked if there is value to conducting research on this topic (n=308, 42.31%), more research funding should be allocated to this field (n=220, 30.26%), and that clinicians should receive training via supplemental education (n= 244, 33.66%) (**Figure 7**). Additionally, most respondents disagreed when asked if they would be comfortable counselling their patients on this topic (n=104, 28.89%), and strongly disagreed when asked if they would be comfortable recommending this therapy to patients (n=123, 34.17%).

**Figure 7.**
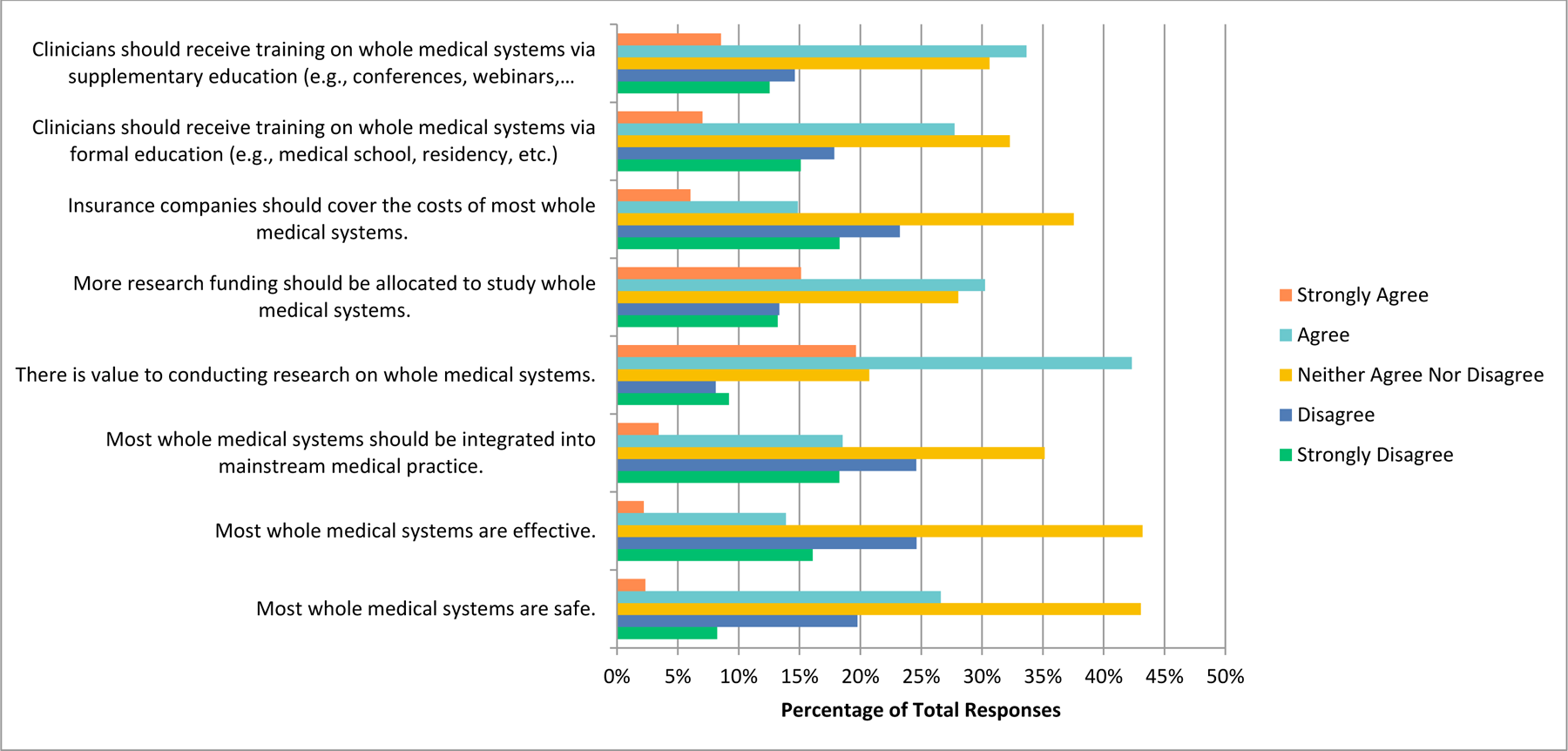
Agreement with the Following Statements Regarding Whole Medical Systems.

### Benefits and Challenges

Participants were also asked which benefits they see associated with CAIM, to which they answered ‘expanded treatment options for patients’ (n=502, 69.53%), ‘focus on prevention and lifestyle changes’ (n=479, 66.34%), ‘holistic approach to health and wellness’ (n=465, 64.40%), ‘cultural and spiritual relevance for certain populations’ (n=423, 58.59%) (**Figure 8**). On the other hand, when asked which challenges they see with CAIM, participants were concerned about ‘lack of scientific evidence for safety and efficacy’ (n=688, 92.97%), ‘lack of standardization in product quality and dosing’ (n=n=657, 88.78%), ‘difficulty in distinguishing legitimate practices from scams or fraudulent claims’ (n=570, 77.03%), and ‘limited regular and oversight’ (n=565, 76.35%) (**Figure 9**).

**Figure 8.**
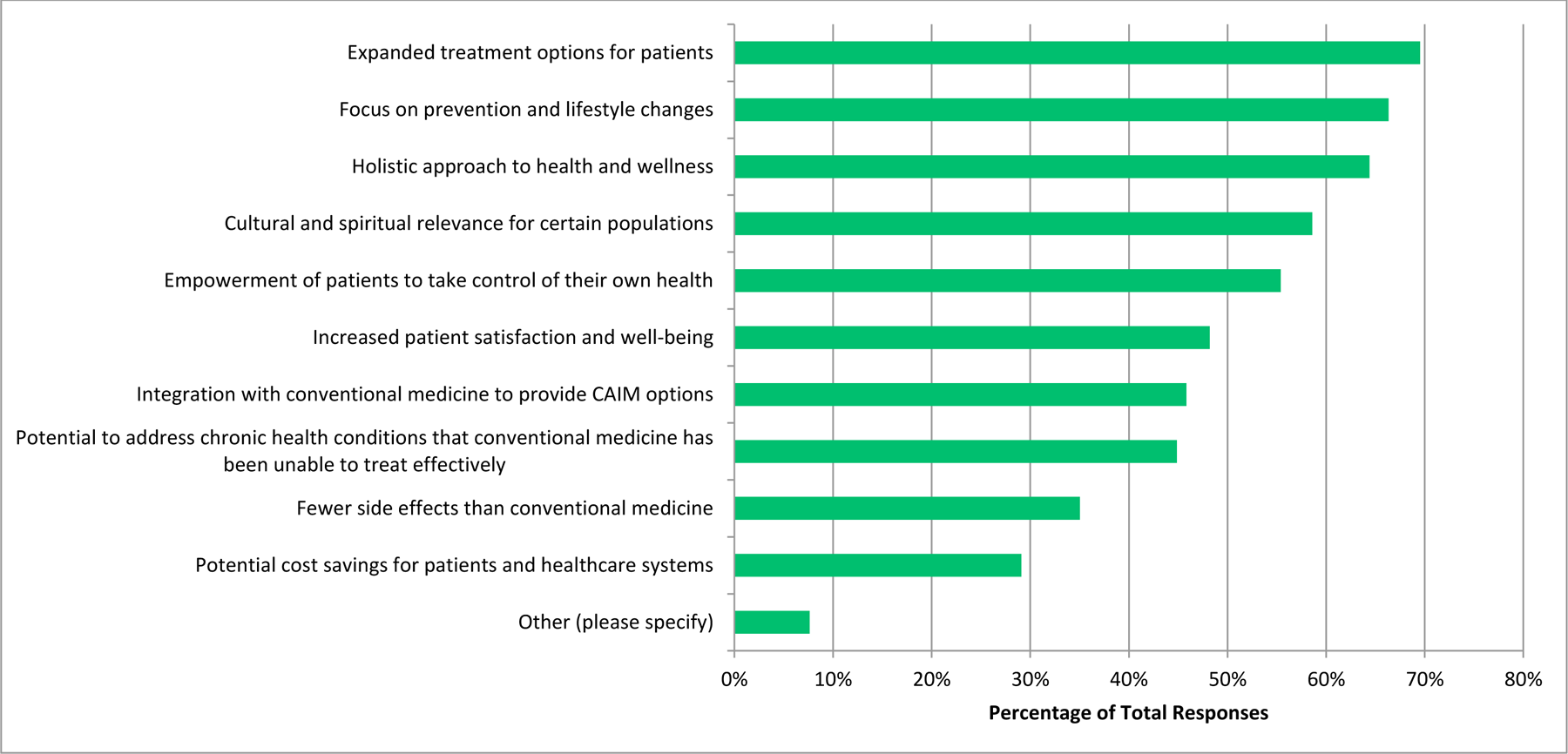
Benefits Perceived to be Associated With CAIM.

**Figure 9.**
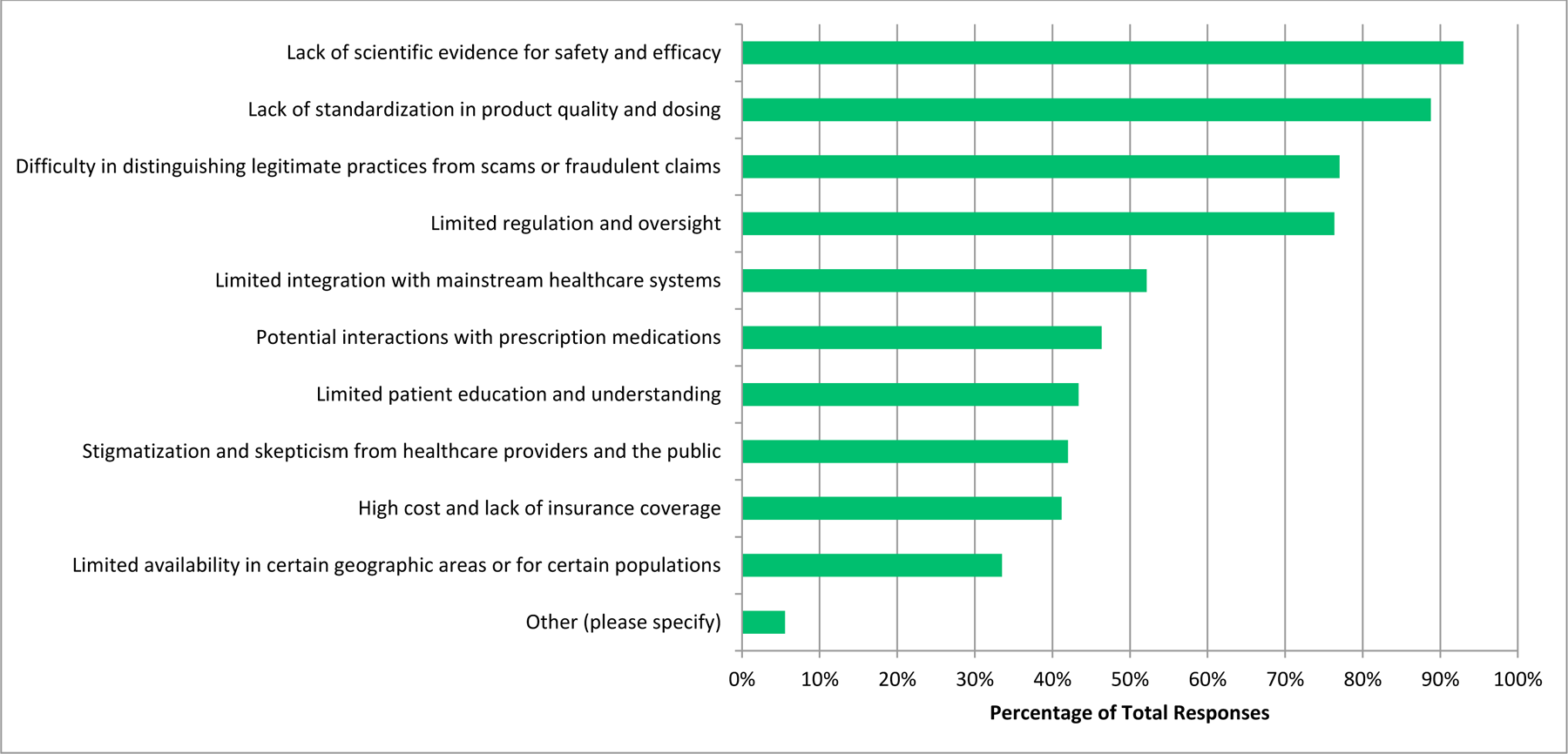
Challenges Perceived to be Associated With CAIM.

### Thematic Analysis

In total, 39 codes were identified from 145 open-ended responses. These codes were then summarized and grouped further to create 5 distinct themes – which are specific patterns that were found in this dataset. Firstly, “polarized opinions on CAIM” included those who were in support, and those that were against the use of different forms of CAIM, including which aspects should receive more attention. Secondly, “concerned about the safety of CAIM practices and the lack of research” encompassed the opinions of many respondents who believe that there should be more research done in the field of CAIM, as it may be dangerous for patients. Next, “integrated approach including CAIM is optimal” included responses that encouraged a combination of different forms of medicine for overall patient wellbeing. Additionally, “proposing alternative research questions” was a theme that many responses were categorized into, as a number of respondents believed that there are different avenues to approach in this area of research. Finally, “pharmaceutical and financial components of healthcare should be considered when investigating CAIM” included those responses which were contrasting CAIM to major pharmaceuticals, and displaying concerns regarding the profit aspect of healthcare. Coding and thematic analysis data are available at: https://osf.io/au7zb.

## Discussion

The objective of this study was to identify the perceptions of psychiatry researchers and clinicians on CAIM. Our findings demonstrate that researchers and clinicians working in the field of psychiatry have positive perceptions towards certain CAIM categories such as mind-body therapies, however they have hesitancies towards others such as whole medical systems. Participants believed that there is value in conducting more research in the field of CAIM, and that health care practitioners should receive educational training on certain CAIM therapies.

These findings are consistent with existing literature that has investigated the perceptions of CAIM among healthcare professionals. Specifically, previous studies have found that therapies such as yoga and tai-chi tend to be the most well-known and recommended CAIM therapies among a cohort of rehabilitation physicians [26], and that a large proportion of physicians would like to receive more training on CAIM [27]. Also, specific to the field of psychiatry, a previous study conducted found that psychologists/psychiatrists have a statistically significant difference in their perceptions of CAIM in comparison to other professionals such as theologists when asked if “nutritional counselling and dietary food/supplements can be effective in the treatment of pathology,” with the theologists being stronger advocates for CAIM [28]. Similarly, a prior study found that while psychologists were open to using CAIM with patients, there was also scepticism surrounding certain aspects [29]. Also, a study found that within a sample of clinical psychologists, there was low knowledge of CAIM, but positive attitudes towards its patient use overall among the respondents [30]. Finally, in a study that compared the beliefs of practicing psychologists with psychology students, the psychologists expressed a greater concern in integrating CAIM therapies [31].

In this study, respondents were presented with a number of CAIM therapies. Overall, mind-body therapies such as meditation, yoga, and tai-chi were considered to be the most promising with regards to prevention, treatment and management of disease, as well as the one therapy group which they seem to consistently recommend to their patients. In addition, respondents agreed that these are safe therapeutic approaches that they feel comfortable counselling and recommending to their patients. Numerous studies that have been conducted have found that yoga is an effective mechanism of improving psychiatric symptoms [32–33], as well as the importance of meditation as an effective coping mechanism [34]. As there are no unknown substances being consumed or used with mind-body therapies, this may explain why respondents feel that it is not harmful to patients in the same way as other CAIM techniques.

Contrarily, whole medical systems, which includes therapies such as Ayurvedic medicine, traditional Chinese medicine, and homeopathy, received more negative responses in this study. Specifically, respondents seem unsure that these practices are safe and effective, and would not feel comfortable in recommending or counselling these services to patients. These practices have been found to be popular in psychiatry in countries such as India and China, which may be attributed to the lack of adequately trained psychiatric personnel to manage the amounts of psychiatric patients [35]. As the majority of respondents from our study were geographically located in the Americas and Europe as opposed to Asia, this may be the reason why they are more reluctant to trust these types of therapies.

When respondents were asked about the various therapies and whether they believe there is value in conducting more research, and if they should receive education; either formally or supplementary in the respective fields, there was a consistently positive response noted. This is consistent with existing literature, as a majority of family health strategy doctors and nurses have also shown an interest in CAIM training [36]. The majority of respondents in the study identified as a researcher, or a researcher and a clinician, which can explain why they believe that doing additional research may be beneficial to this area. Moreover, many respondents in the study identified as being in a later stage in their career (faculty members, principal investigators, senior researchers), which could explain why they believe that receiving education on these topics is important. If they had been provided this education earlier, they may be more equipped to counsel and recommend these therapies to patients. Finally, from the thematic analysis conducted, it was evident that respondents believe that there is value in conducting research in this field, as “Proposing alternative research questions/populations/approach” was the theme that contained the most coded responses.

### Strengths and Limitations

In terms of study strengths, this study was able to generalize psychiatry researcher and clinician perceptions of CAIM, as the survey included a large and international sample of individuals who had varying opinions on the topic. In addition, emails were sent to individuals who have published in recent years, thus reducing the proportion of invalid and inactive email addresses. Additionally, multiple reminder emails were sent in order to increase the response rate. Conversely, some limitations to this study include that it is most likely that we only sampled participants who understand English, as only journals that publish in English are being included and the survey was in English. Additionally, due to the nature of the sampling strategy, which includes extracting the name and email address of the corresponding author of articles published in scholarly journals, there were proportionately more researchers than clinicians being contacted. Another important limitation that must be considered is that CAIM is an umbrella term, and although we have separated it into 5 categories (i.e. mind body therapies, biologically based therapies, manipulative and body-based practices, biofield therapies, and whole medical systems), the safety and efficacy profiles for each therapy differ [37]. Therefore, participants had to generalize their thoughts on these therapies, as opposed to provide their individual thoughts on each respective type. The response rate is also anticipated to be underestimated due to the fact that some email addresses were inactive or invalid; furthermore, some contacted individuals were retired, ill, have passed away, or were otherwise unable to respond for other reasons. Next, there is the possibility of individuals who received the invitation and choose not to take the survey. As a result, non-response bias, which occurs when there is a difference in characteristics between the responders and non-responders to the survey [38] may be present in the study, meaning the results may not be as representative of the population. Finally, there is also potential for recall bias, which occurs when participants have differing recall of information [38].

## Conclusions

In this study, we attempted to gain an understanding on the perceptions of psychiatry researchers and clinicians on various CAIM modalities. Respondents were given the option to rank their opinions on a number of therapies, and provide additional information that they feel should be considered. This provided valuable insight on the current perceptions of CAIM in a growing medical field such as psychiatry, and can form the basis for further research to be conducted, as the respondents feel strongly that this should be done. These findings can also be used to develop tailored educational resources, as respondents agree that this would be beneficial in their training. Previous literature has suggested that CAIM interventions can improve psychiatric conditions, however this is the first study to explore this specific research question and to gain an understanding on psychiatry researcher and clinician perceptions. It is hoped that these findings and the analysis can contribute to the field of psychiatry as well as the field of CAIM, such that it can provide a basis for further research to follow.

## Data Availability

All data and materials associated with this study has been posted on the Open Science Framework.

https://doi.org/10.17605/OSF.IO/W6HNM

## List of Abbreviations

CAIM: complementary, alternative, and integrative medicine
DSM-5: Diagnostic and Statistical Manual of Mental Disorders-5
HIV: Human Immunodeficiency Virus
ICD-11: International Classification of Diseases-11
SAM-e: S-adenosylmethionine

## Declarations

### Ethics Approval and Consent to Participate

This study received approval from the University Tübingen Research Ethics board (REB Number: 389/2023BO2).

### Consent for Publication

All authors consent to this manuscript’s publication.

## Availability of Data and Materials

All data and materials associated with this study has been posted on the Open Science Framework and can be found here: <u>10.17605/OSF.IO/W6HNM</u>

## Competing Interests

The authors declare that they have no competing interests.

## Funding

This study was unfunded.

## Authors’ Contributions

JYN: designed and conceptualized the study, collected and analysed data, drafted the manuscript, and gave final approval of the version to be published.

JK: assisted with the collection and analysis of data, made critical revisions to the manuscript, and gave final approval of the version to be published.

HC: assisted with the design and concept of the study and the analysis of data, made critical revisions to the manuscript, and gave final approval of the version to be published.

## Notes

### Competing Interest Statement

The authors have declared no competing interest.

### Clinical Protocols

https://doi.org/10.17605/OSF.IO/75ACV

### Author Declarations

This study received approval from the University Tubingen Research Ethics board (REB Number: 389/2023BO2).

